# The evaluation of a novel tool to remotely assess visual acuity in chronic uveitis patients during the COVID-19 pandemic

**DOI:** 10.1101/2021.04.14.21255457

**Authors:** J.L.J. Claessens, J. van Egmond, J.H. de Boer, R.P.L. Wisse

**Affiliations:** Department of Ophthalmology, University Medical Center Utrecht, Utrecht, the Netherlands; Faculty of Medicine, Utrecht University, Utrecht, the Netherlands

## Abstract

**Background:** Restrictions due to the recent COVID-19 pandemic catalysed the deployment of telehealth solutions. A novel web‐based visual acuity test, validated in a healthy population, may be of great value in the follow‐up of uveitis patients.

**Objective:** To determine the measurement accuracy of the unsupervised remote Easee web‐based visual acuity test in uveitis patients, when compared to a conventional in‐hospital assessment.

**Methods:** Cross‐sectional diagnostic accuracy study. Between April 2020 and September 2020, consecutive adult uveitis patients were invited for the web‐based visual acuity test (index test) within two weeks prior to their conventional in‐hospital assessment (reference test).

**Results:** A total of 269 patients were invited by mail, of whom 84 visited the website (31%). Ultimately 98 eyes met the criteria for statistical analysis. The mean difference between the two tests was low and non‐significant: 0.02 logMAR (SD 0.12, *P* = 0.085). The 95% limits of agreement ranged from ‐0.21 to 0.26 logMAR. No relevant differences were identified in clinical characteristics between subgroups with a good performance (i.e. difference between the tests ≤0.15 logMAR) or underperformance (i.e. difference >0.15 logMAR) on the web‐based test.

**Conclusion:** The web‐based visual acuity test is a promising tool to remotely assess visual acuity in the majority of uveitis patients, especially relevant when access to ophthalmic care is limited. No association between patient‐ or uveitis‐related variables and (under)performance of the test were identified. These outcomes underline the potential of remote vision testing in other common ophthalmic conditions. A proper implementation of this web‐based tool in health care could be of great value for revolutionizing teleconsultations.

## Introduction

The COVID-19 pandemic impacted almost every aspect of life and put a lot of stress on health care systems. Most elective outpatient care had to be cancelled or rescheduled.^1^ At the ophthalmological department of our academic hospital, the University Medical Centre Utrecht (UMCU), the Netherlands, a telehealth approach was introduced to assist with triaging urgency of appointments.^2^ As such, patients were requested to perform a remote unsupervised visual acuity test, of which the validity was recently investigated by our research team just prior to COVID-19.^3^

Uveitis is an important cause of blindness in both developed (7‐15%) and developing countries (24‐ 25%).^4–6^ It describes a heterogeneous group of inflammatory conditions of the eye with multiple clinical presentations, classified by The Standardization of Uveitis Nomenclatur (SUN) Working Group. & Common symptoms of disease activity include blurred vision, ocular pain, eye redness, and photophobia.^10,11^ Despite therapeutic strategies aiming to reverse visual loss in early disease stages, advanced disease complications, such as macular oedema, glaucoma, or retinal scars, can result in irreversible visual loss.^12,13^ In order to minimise the damage and incidence of complications, early detection and timely treatment are of great importance.^14^ The UMC Utrecht is a Center of Excellence and national referral clinic for the diagnosis and treatment of uveitis.^15^

Web‐based self‐tests can remove barriers to care and have the potential to enrich teleconsultations. Tools for self‐testing of visual acuity these have already been successfully implemented in screening programs of visual impairment.^16,17^ We hypothesize that remote self‐assessment tools could reduce frequencies of hospital visits, thereby lowering costs for patients for travelling, parking, taking time off from work, as well as saving valuable hospital resources and staff. Especially considering the acute reduction in access to health care during the pandemic, telehealth solutions are of great value to continue (eye) care delivery. The Amsterdam‐based med‐tech start‐up company Easee BV, offers the world’s first *Conformité Européenne* (CE) certified web‐based visual acuity test that complies with all relevant International Organization for Standardization (ISO) standards. The accuracy of the web‐based test by Easee BV to assess visual acuity has been validated in healthy individuals^3^ and in keratoconus patients.^18^ Promising results were found, especially in higher visual acuity ranges.^18^ We hypothesize that an unsupervised self‐assessment of visual acuity is a reliable substitute for the conventional in‐hospital assessment, also in more complex patients. Uveitis patients are a challenging domain, since our institution hosts a particularly complex population, with a favourable age distribution regarding digital proficiency.

## Methods

### Study design and recruitment of data

This study was conducted at the University Medical Center Utrecht (UMCU), the Netherlands, a uveitis Center of Excellence and expertise center. Adult uveitis patients were invited by telephone or by mail to do the web‐based test at home, prior to their hospital consultation, between April 2020 and September 2020. For this study, we included the patients who performed the Web‐based visual acuity test one to fourteen days before the conventional in‐hospital visual acuity assessment. Patients were instructed to redo the web‐based eye exam, or reach out to the study team, whenever they experienced a change in visual acuity in the interval between the two assessments. We excluded patients who were unable to complete the Web‐based test successfully, who did not have a reference test outcome assessed within the 14‐days interval, who coincidentally changed their glasses or contact lenses, or who reported a change in subjective visual acuity between the two assessments without redoing the web‐based eye exam.

For each patient, the following data were collected from the electronic health record: sex, age, uveitis classification, medical history, use of medication, symptoms associated with uveitis, any coexisting ophthalmic disease at time of the appointment, and visual acuity outcomes. All ophthalmologists in the UMCU practice according the SUN classification criteria.^9^ Disease activity was classified based on the vitreous haze (VH), anterior chamber cell count (ACC), optical coherence tomography and fluorescent angiography and dichotomized into, ‘inactive’ (ACC and VH ≤ 0.5 and not considered active by the expert) or ‘active’ (ACC or VH > 0.5 or considered as active by the expert).

### Web‐based assessment (index test) and conventional visual acuity assessment (reference test)

Patients were invited to perform the web‐based test in their home environment, prior to their hospital appointment. The test is accessible via a dedicated URL. Users need a computer or tablet, and a smartphone to do the test. In short, the smartphone functions as a remote control by which the user submits input from a distance of 3 meters to the computer screen. The set‐up is depicted in Figure 1 below. The screen displays a sequence of optotypes that the user must correctly identify. Audio instructions guide the user through the test. Patients are asked to wear their habitual spectacles or contact lenses, if applicable. Patients can choose to measure one or both eyes. We instructed patients to assess both.

**Figure 1:**
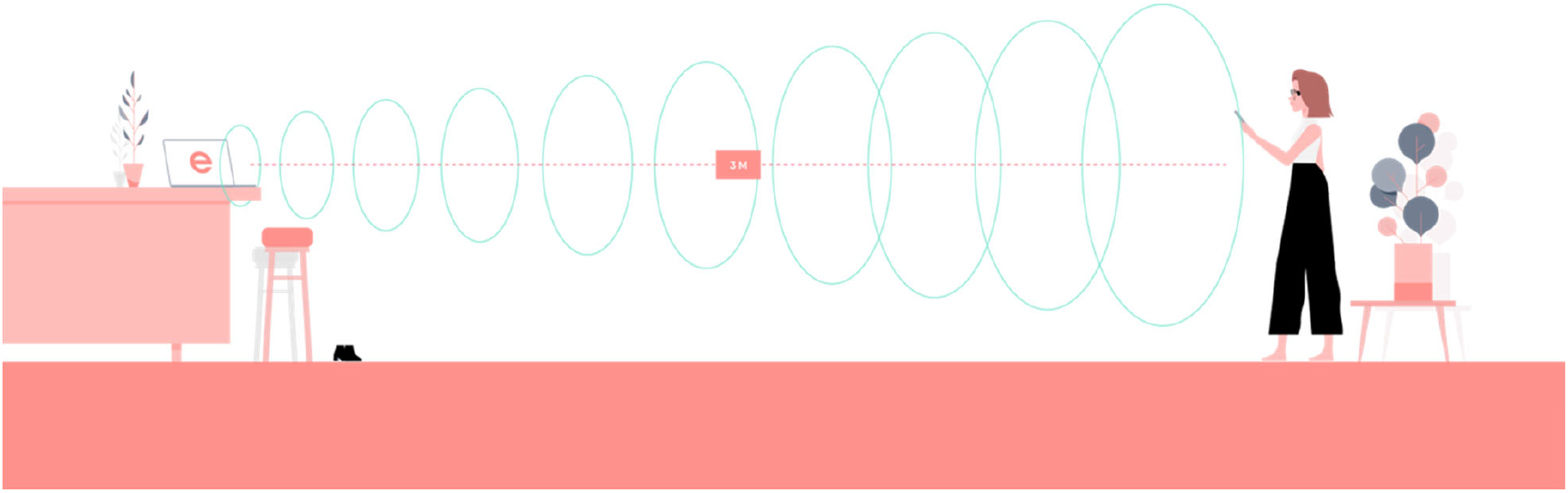
Set‐up of the web‐based test by Easee B.V.

Conventional measurements were assessed during routine consultations by ophthalmologists, residents in training, or optometrists, using a Snellen chart (the gold standard in our hospital). The health care providers were blinded for the previous web‐based test results of the subjects.

### Statistical analysis

Our main outcome is the measurement accuracy of the web‐based visual acuity test, as compared to the conventional visual acuity assessment at hospital consultations within 14 days afterwards. The web‐based outcome was reported in logMAR. The Snellen decimal score of the hospital assessments was converted to the logarithm of the minimum angle of resolution (logMAR). The measurement accuracy is expressed by mean differences between the two assessments, and 95% Limits of Agreement (95%LoA). The mean difference can be interpreted as the systematic difference between the assessments (bias) and the 95%LoA as the range within 95% of the differences between one assessment and the other are included (random error). This methodology was first introduced by Bland and Altman, and is commonly used in method comparison studies to evaluate the agreement between two measurements on a continuous scale.^19^

Variable outcomes when repeatedly performing visual acuity tests in an individual are common.^20–22^ We will consider a difference of more than ±0.15 logMAR clinically relevant. This is supported by an authorative cross‐sectional study in a large eye clinic using various charts and observers.^20^ The minimum visual acuity (VA) score that can be determined by the web‐based test is 0.05 Snellen decimal (1.3 logMAR). Lower scores will be described as “<0.05 Snellen decimal” (>1.3 logMAR). As their exact test value was unknown, patients with these poor VA scores were not compared in the Bland‐Altman analysis, but descriptively analysed as a subgroup.

We did an additional subgroup analysis to investigate associations between clinical characteristics and the agreement between the index and reference test: patients with differences greater than 0.15 logMAR (i.e. underperformance of the web‐based test), vs. patients with differences smaller than or equal to 0.15 logMAR (i.e. good performance of the web‐based test). Chi‐squared and independent sample t‐tests were used to assess differences between these groups. To correct for bilaterality (both eyes of the same patient included), the statistical tests for age, sex, and interval between the two tests, were presented per eye separately.

A multivariate generalized estimating equation (GEE) assessed the association between independent variables and visual acuity outcomes of both tests. The GEE model was constructed to correct for bilaterality, age, sex, use of mydriatics, visual acuity influencing comorbidities, symptoms associated with uveitis activity and the interval in days between the two assessments.

### Ethical considerations

The study was performed in accordance with Dutch privacy laws and the Declaration of Helsinki. The data collected by Easee BV is stored on GDPR and HIPAA compliant servers in the EU. All included patients were registered in the registry ‘Ocular inflammation, METC 17‐363’, that contains a written informed consent for the use of data.

## Results

### Participation

A total of 269 patients were invited by mail to execute the web‐based exam at home, prior to their hospital visit. Ultimately, 84 patients visited the Easee website, and 72 patients finished the test successfully. Participants did not structurally report their reason for non‐participation, though. Patients who visited the URL were significantly younger than those who did not (47.2 vs. 53.3 years, p=0.012). The participation flowchart can be found in the supplemental data.

### Included patients

A total of 59 patients met all inclusion criteria, and 98 eyes were included for analysis (not all patients followed the instruction to assess both eyes). The clinical characteristics of the study population are summarized in Table 1. Most of the included patients were female (67.8%). The mean interval between the two tests was 4.8 days ±2.7 days. At time of the appointment, symptoms of potentially active uveitis, such as ocular pain, floaters, photophobia, or visual loss, were present in 26.5% of the eyes. Based on the SUN classification^9^ only 27.1% of the patients had uveitis anterior and 96.6% had a chronic course. At time of the appointment, 24.5% of the eyes were described as “active inflammation” of uveitis, while 74.5% of the eyes were “inactive”.

**Table 1.** Clinical characteristics of the study population

| Clinical characteristics |  | n = 59 patients |
| --- | --- | --- |
| Age (years), mean (SD) |  | 46.5 (15.1) |
| Sex | Male | 19 (32.2%) |
|  | Female | 40 (67.8%) |
| Interval between tests (days), mean (SD) |  | 4.8 (2.7) |
| Ophthalmic medication<sup>a</sup> | Mydriatics | 4 (6.8%) |
|  | Other | 45 (76.3%) |
| Uni- or bilateral uveitis | Unilateral | 16 (27.1%) |
|  | Bilateral | 43 (72.9%) |
| Anatomical classification<sup>b</sup> | Anterior | 16 (27.1%) |
|  | Non-anterior | 43 (72.9%) |
| Uveitis course<sup>b</sup> | Acute | 2 (3.4%) |
|  | Chronic | 57 (96.6%) |
| Uveitis characteristics (per eye) |  | n = 98 eyes |
| Activity of uveitis | Inactive<sup>c</sup> | 73 (74.5%) |
|  | Active<sup>d</sup> | 24 (24.5%) |
|  | Not known | 1 (1.0%) |
| Visual acuity influencing comorbidities, at time of appointment<sup>e</sup> |  | 30 (30.6%) |
| Anamnestic symptoms of uveitis<sup>f</sup> |  | 26 (26.5%) |
| All values are reported as: n (%), unless otherwise specified.<br>a: use of ophthalmic medication at time of the in-hospital appointment<br>b: according the SUN classification<sup>9</sup><br>c: when ACC and VH is $\leq 0.5$ and not called active by the ophthalmologist<br>d: when ACC or VH is $\geq 1$ or called active by ophthalmologist<br>e: including: (secondary) cataract, keratitis, scleritis, corneal lesion or history of pars plana vitrectomy<br>f: symptoms associated with uveitis: ocular pain, floaters, photophobia, visual loss |  |  |

### Measurement accuracy of the Web‐based visual acuity test

The mean web‐based VA was 0.12±0.25 logMAR (0.86±0.37 Snellen), compared to 0.10±0.25 logMAR (0.89±0.32 Snellen) for the conventional Snellen chart. The Bland‐Altman plot in Figure 2 visualizes the comparisons between the two tests for 91 comparisons (seven patients had a web‐based test‐score of <0.05 Snellen). The mean difference was 0.02 ±0.12 logMAR (p=0.085). The 95%LoA ranged from ‐0.21 to 0.26 logMAR, without indications for a proportional bias. Overall, a majority of comparisons (76.9%, n=70) fell within the predetermined accepted deviation limit of ±0.15 logMAR. The cumulative distribution of the differences demonstrates that most of the differences were close to zero (i.e. no difference).

**Figure 2.**
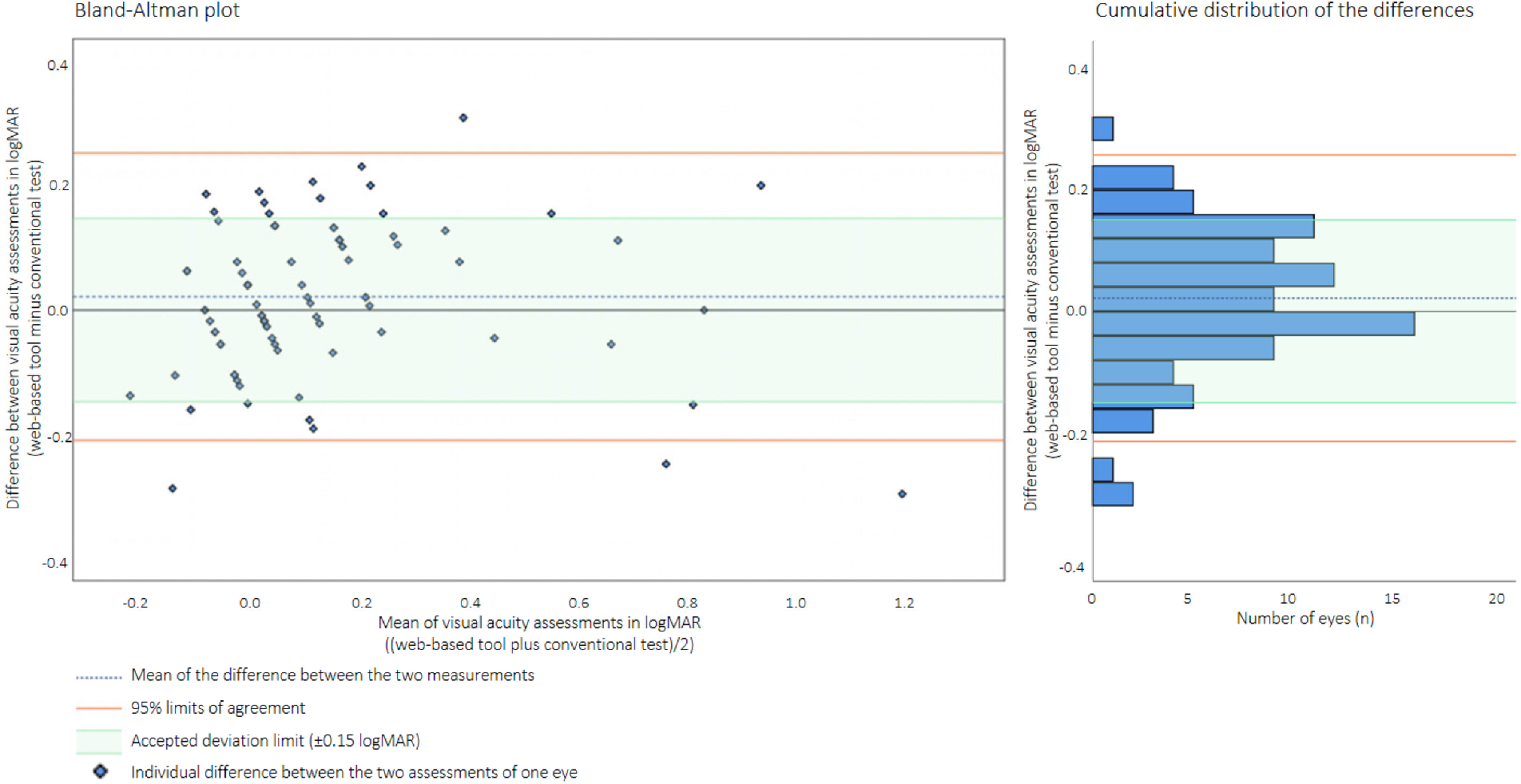
The comparisons of the web‐based tool vs. the conventional test. Left: Bland‐Altman plot depicting the differences of the VA assessments in logMAR. Individual differences between the two assessments are plotted per eye. Right: Cumulative distribution of the VA differences in logMAR.

### Subgroup analysis of poor visual acuity outcomes

The visual acuity scores of seven patients were lower than the minimum detectable value of the web‐based test. For six of these patients, the visual acuity was correctly determined by the web‐based test: these patients also had a visual acuity test score of <0.05 Snellen on the conventional assessment. Surprisingly, one patient had a reference test score of 0.4 Snellen decimal. Upon inquiry, the patient reported that the test was technically difficult to perform, and that the outcome therefore was not representative for the actual visual acuity status.

### Subgroup analysis of good vs. under performance of web‐based test

The results of the subgroup analysis are shown in Table 2. Multiple trends were visible in the ‘underperformance’ group (n=21 eyes) when compared to the ‘good performance’ group (n=70 eyes), though none of these differences reached statistical significance. In the ‘underperformance’ subgroup, mean age was higher (for OS: 50.67 vs. 43.89 years, p=0.250). Also, there were more visual acuity influencing comorbidities at the time of appointment (42.9% vs. 24.3%, p=0.098), and less anterior uveitis (14.3% compared to 34.3%, p=0.078) in the eyes that showed poorer agreement between two tests. Furthermore, a higher percentage of inactive uveitis (85.7% compared to 71.0%, p=0.176) was seen in the underperformance group.

**Table 2.**
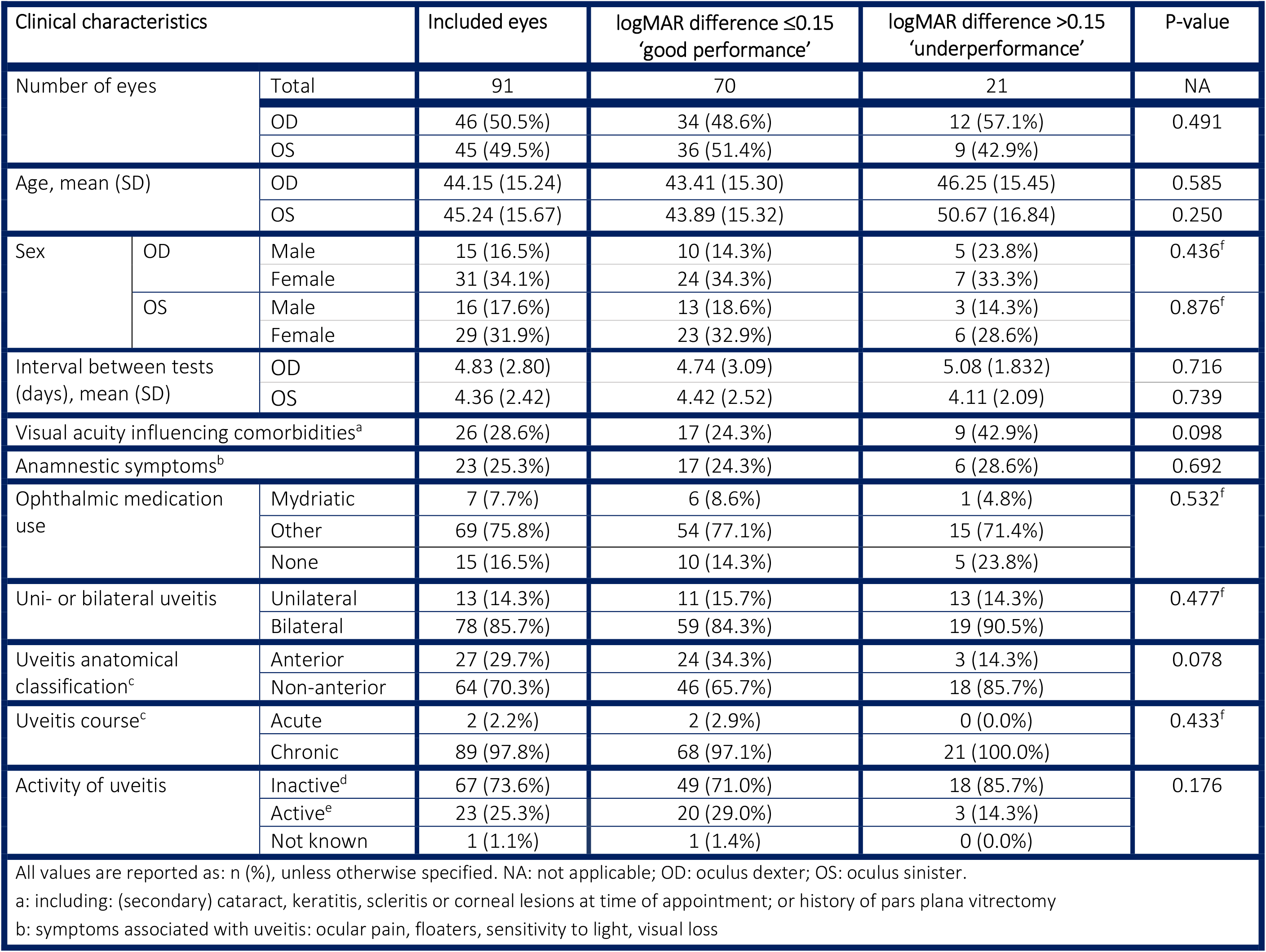
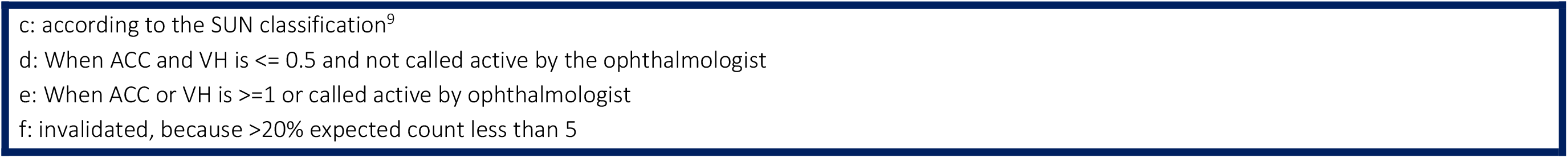
Subgroup analysis of “good performance” vs. “underperformance” of Web‐based test

| Clinical characteristics |  |  | Included eyes | logMAR difference ≤0.15<br>‘good performance’ | logMAR difference >0.15<br>‘underperformance’ | P-value |
| --- | --- | --- | --- | --- | --- | --- |
| Number of eyes |  | Total | 91 | 70 | 21 | NA |
|  |  | OD | 46 (50.5%) | 34 (48.6%) | 12 (57.1%) | 0.491 |
|  |  | OS | 45 (49.5%) | 36 (51.4%) | 9 (42.9%) |  |
| Age, mean (SD) |  | OD | 44.15 (15.24) | 43.41 (15.30) | 46.25 (15.45) | 0.585 |
|  |  | OS | 45.24 (15.67) | 43.89 (15.32) | 50.67 (16.84) | 0.250 |
| Sex | OD | Male | 15 (16.5%) | 10 (14.3%) | 5 (23.8%) | 0.436<sup>f</sup> |
|  |  | Female | 31 (34.1%) | 24 (34.3%) | 7 (33.3%) |  |
|  | OS | Male | 16 (17.6%) | 13 (18.6%) | 3 (14.3%) | 0.876<sup>f</sup> |
|  |  | Female | 29 (31.9%) | 23 (32.9%) | 6 (28.6%) |  |
| Interval between tests (days), mean (SD) |  | OD | 4.83 (2.80) | 4.74 (3.09) | 5.08 (1.832) | 0.716 |
|  |  | OS | 4.36 (2.42) | 4.42 (2.52) | 4.11 (2.09) | 0.739 |
| Visual acuity influencing comorbidities<sup>a</sup> |  |  | 26 (28.6%) | 17 (24.3%) | 9 (42.9%) | 0.098 |
| Anamnestic symptoms<sup>b</sup> |  |  | 23 (25.3%) | 17 (24.3%) | 6 (28.6%) | 0.692 |
| Ophthalmic medication use |  | Mydriatic | 7 (7.7%) | 6 (8.6%) | 1 (4.8%) | 0.532<sup>f</sup> |
|  |  | Other | 69 (75.8%) | 54 (77.1%) | 15 (71.4%) |  |
|  |  | None | 15 (16.5%) | 10 (14.3%) | 5 (23.8%) |  |
| Uni- or bilateral uveitis |  | Unilateral | 13 (14.3%) | 11 (15.7%) | 13 (14.3%) | 0.477<sup>f</sup> |
|  |  | Bilateral | 78 (85.7%) | 59 (84.3%) | 19 (90.5%) |  |
| Uveitis anatomical classification<sup>c</sup> |  | Anterior | 27 (29.7%) | 24 (34.3%) | 3 (14.3%) | 0.078 |
|  |  | Non-anterior | 64 (70.3%) | 46 (65.7%) | 18 (85.7%) |  |
| Uveitis course<sup>c</sup> |  | Acute | 2 (2.2%) | 2 (2.9%) | 0 (0.0%) | 0.433<sup>f</sup> |
|  |  | Chronic | 89 (97.8%) | 68 (97.1%) | 21 (100.0%) |  |
| Activity of uveitis |  | Inactive<sup>d</sup> | 67 (73.6%) | 49 (71.0%) | 18 (85.7%) | 0.176 |
|  |  | Active<sup>e</sup> | 23 (25.3%) | 20 (29.0%) | 3 (14.3%) |  |
|  |  | Not known | 1 (1.1%) | 1 (1.4%) | 0 (0.0%) |  |
| All values are reported as: n (%), unless otherwise specified. NA: not applicable; OD: oculus dexter; OS: oculus sinister. |  |  |  |  |  |  |
| a: including: (secondary) cataract, keratitis, scleritis or corneal lesions at time of appointment; or history of pars plana vitrectomy |  |  |  |  |  |  |
| b: symptoms associated with uveitis: ocular pain, floaters, sensitivity to light, visual loss |  |  |  |  |  |  |

c: according to the SUN classification<sup>9</sup>
d: When ACC and VH is $\leq 0.5$ and not called active by the ophthalmologist
e: When ACC or VH is $\geq 1$ or called active by ophthalmologist
f: invalidated, because $>20\%$ expected count less than 5

### Generalized estimating equation analysis

The GEE analysis revealed no significant associations between the independent variables and the visual acuity outcomes of the tests. There are no factors that seem to influence visual acuity outcomes of both tests individually, or the difference between the two tests. The studied associations and their outcomes can be found in the Supplemental data.

## Discussion

This study determined the diagnostic accuracy of the novel Easee web‐based test to remotely self‐assess visual acuity in complex uveitis patients, and compared these outcomes to a conventional in‐hospital assessments shortly afterwards. Based on our findings, the web‐based test seems a promising instrument to estimate visual acuity of uveitis patients in their home‐environment, especially relevant when access to health clinics is limited. We found a clinically negligible mean difference of 0.02 ±0.12 logMAR between the web‐based test and the conventional chart assessment. This mean difference is evidently lower than the predetermined accepted deviation limit of ±0.15 logMAR. The distribution of the differences, expressed by the 95%LoA, ranged from ‐0.21 to 0.26 logMAR, which slightly exceeds the predetermined accepted deviation limit, meaning that some of the individuals had higher differences between the two assessments than expected based on normal measurement variation. Notwithstanding, the agreement between the two assessments was good for the majority of the comparisons (77%). Sub‐group analyses identified no significantly clinical or patient factors influencing the agreement between the two tests.

When comparing visual acuity assessments, there are two important phenomena that should be taken into account. First, there is always variation when repeatedly assessing visual acuity within the same individual, even in the absence of a clinical change. A study that focused on test‐retest variability of Snellen visual acuity charts reported 95%LoA ranging from ±0.18 logMAR (single letter method) to ±0.33 logMAR (line assignment method).^21^ The line assignment method of the Snellen chart (where the test is terminated when half or more of the letters are misread) remains the most popular method in clinical practice, despite the introduction of more reliable alternatives such as the ETDRS chart^22^ (with reported test‐retest variabilities ranging from ±0.07 to ±0.18 logMAR^23,24^). Convenience (familiarity, short test duration) seems to outweigh accuracy in large and busy eye clinics. When comparing the abovementioned test‐retest 95%LoA to the range found in our study, the remote web‐based exam does not appear to underperform compared to the Snellen chart’s line assignment method, the most used VA test in our hospital.

A second phenomenon, is that there is always variation when two different types of visual acuity charts are compared.^20,25,26^ The web‐based test used a presentation of optotypes based on tumbling‐E and proprietary optotypes, while the conventional exam was a Snellen letter chart. A conversion effect contributes partly to the observed differences in outcomes between the Web‐based test and the conventional chart.

Before this study, our team had evaluated the accuracy of the web‐based eye exam of Easee in a cohort of healthy individuals^3^ and a cohort of keratoconus patients.^18^ When comparing our current results to previous findings, the measurement accuracy of the tool appears to be improved: while the mean differences in VA were already comparable, the current distribution of VA differences is evidently smaller (e.g. in healthy individuals the 95%LoA were ±0.43 logMAR from the mean difference). Clearly, the measurement accuracy has been improved as the algorithm and test flow of the tool have been updated over time. An important difference with the previous studies, however, is that in this study we focused on corrected visual acuity, whilst previously, uncorrected visual acuities were assessed. For uncorrected VA assessments, there will be individuals in the lower VA ranges due to unmitigated refractive errors. Accuracy of assessments is known to be suboptimal in these lower VA ranges, especially for Snellen charts.^22^ In higher visual acuity ranges, web‐based tools appear very reliable, as demonstrated by this study.

This was the first study from our group to evaluate the web‐based tool assessments in a completely unsupervised condition; in patient’s own home environment, where lightning and test set‐up were only controlled by the algorithm itself. We value this greatly, as it best reflects the true observations after implementing these tools in clinical practice. Notably, the unsupervised conditions did not seem to materially impact diagnostic accuracy.

A limitation of the study design was that only patients were included who were willing to participate, and who were able to successfully complete the web‐based eye exam in their home environment. The study design induced a participation bias of digitally competent patients, often of a younger age, which might affect the generalizability of the study outcomes. Diagnostic accuracy might be poorer in less digitally competent patients, as remote exams might be performed incorrectly. However, it is important to bear in mind that our inclusion criterion of successful completion of the web‐based eye exam does not imply adequate performance. In other words: a patient can ‘finish’ an exam (at home) without conducting it correctly. In this study, these patients were included in our analyses as well, as information of outliers is important for interpreting the measurement accuracy and identifying patient characteristics that relate to bad performance.

Another possible limitation of the design is the interval between the two assessments. Ideally, two compared VA assessments should be conducted within a time interval as short as possible, to prevent clinical changes to impact observed differences. We consider the mean interval in our interval to be fairly short (mean 5 days ± 3 days). In addition, patients were instructed to redo the web‐based exam or contact the research team if they experienced a change in visual acuity between performing the web‐based eye exam and their hospital appointment. We therefore do not expect the interval to have strongly impacted the observed differences.

## Future perspectives

The results of this study indicate that an unsupervised remote web‐based visual acuity test is a valid replacement of an in‐hospital assessment for the patients who are willing and able to perform a self‐test. It has the potential to enhance teleconsultations in ophthalmic care and creates opportunities for chronic uveitis patients to self‐assess their visual acuity at home, when they are suspecting that their visual function deteriorates. From our own patient board we learned that, this self‐control is considered very important for reassurance and psychological comfort. Most patients seem to be welcoming this tool. Potentially, a web‐based eye exam in combination with telephonic‐ or video‐consultations, might increase future intervals between periodic follow‐up measurements in chronic (and mostly stable) patients, reducing the burden and costs of frequent hospital visits. We strongly recommend introducing telehealth tools like these on individual levels and in close dialogue with the patient. Personalised risk analyses and adequate patient‐instructions are essential. We do not claim this web‐based tool to be a replacement of an ophthalmic investigation nor that it is by itself sufficient for adequate follow‐up in all uveitis patients. A remote web‐based eye exam will obviously never replace complete ophthalmic examinations, including split lamp examinations, essential for cell counting. Importantly, not all uveitis patients will notice visual loss a first symptom loss of increasing activity. ^27–29^ Based on our subgroup analyses, it is not possible to preselect uveitis patients for whom this web‐based test is considered unsuitable. The sample sizes of the subgroups were small, so true effects might have been missed due to a lack of power. We hypothesize that the test performance will be mostly influenced by individual factors such as digital competence or environmental test conditions (such as lighting and test set‐up).

In general, we expect remote visual acuity assessments to be universally applicable for both healthy individuals, as well as individuals with ophthalmic conditions. We propose that the measurement accuracy that we found in this study will not be materially different for similar‐aged patients with eye conditions other than uveitis. An interesting avenue would be to enrich the web‐based platform with a near vision test (reading distance) to internally validate the VA results. Another interesting feature would be the remote assessment of contrast sensitivity and stray light, since both are related to quality of vision and might influence the visual acuity measurements. Unsupervised telehealth tools require a certain level of digital skills of the intended user. We assume that a lower digital competence influences the willingness to perform the test and that it might impact the diagnostic accuracy of unsupervised remote exams (as exams might be performed incorrectly). We recommend future usability testing and clinical research to focus the performance of the web‐based tool in older patients, prior to implementation of these tools in clinical practice.

## Conclusion

The web‐based Easee visual acuity test shows a reliable diagnostic accuracy in the large majority of the studied uveitis patients. No uveitis‐related factors influencing accuracy of the web‐based test were identified. A proper implementation of this web‐based tool in health care could be of great value for enriching teleconsultations, especially when access to hospital care is limited.

## Data Availability

The data are not publicly available, but are available from the corresponding author on reasonable request.

## Supplemental data

**Supplemental Figure 1.**
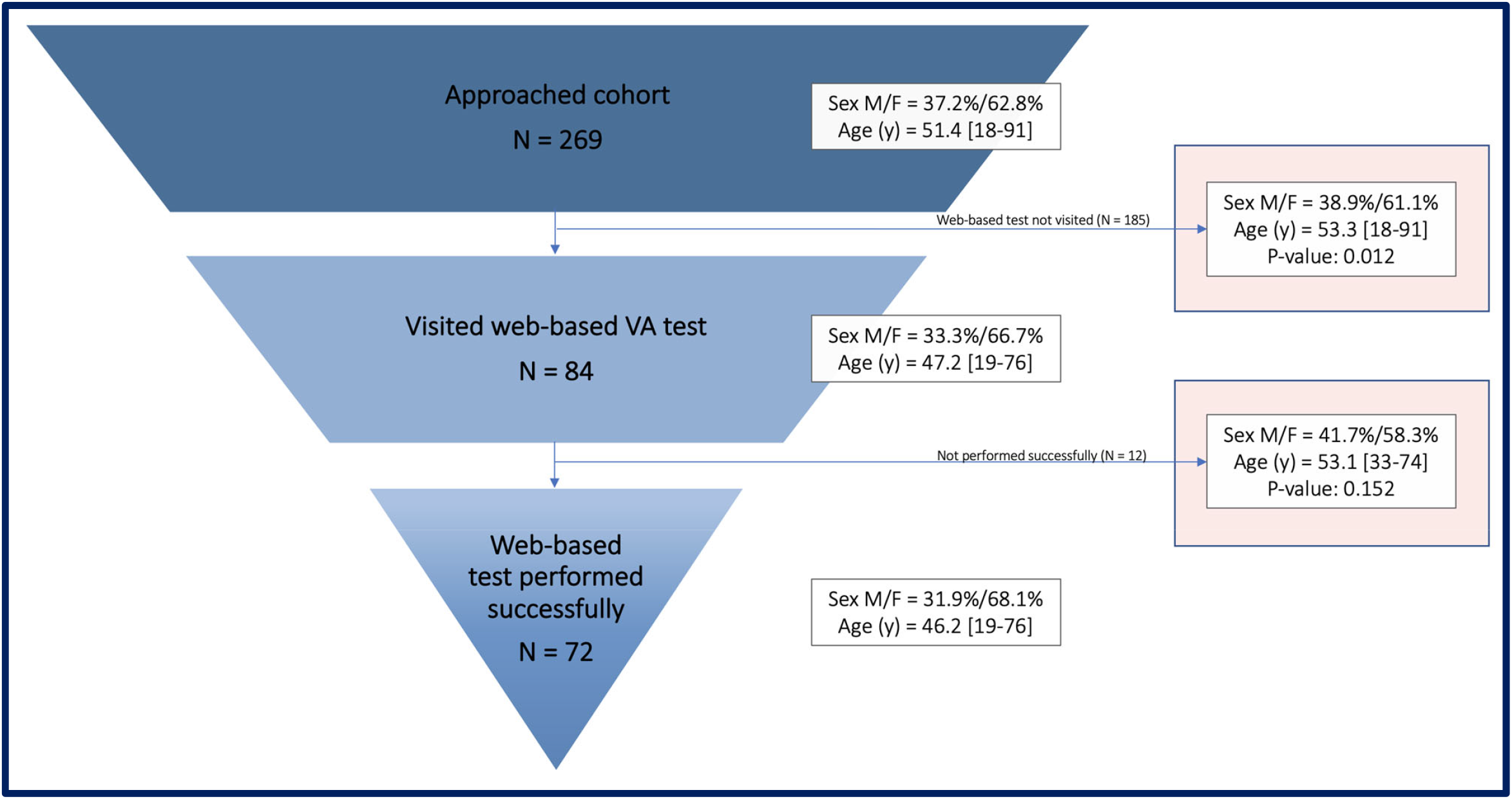
Flow‐chart of participation

**Supplemental Table 1.** GEE analysis

|  |  |  |  |  |  |  |  |  |  |
| --- | --- | --- | --- | --- | --- | --- | --- | --- | --- |
| Multivariate analysis to identify the associations between independent variables and the visual acuity outcome for the difference in logMAR between the web-based BCVA and manifest BCVA, the web-based BCVA in logMAR and the manifest BCVA in logMAR. |  |  |  |  |  |  |  |  |  |
|  | LogMAR difference between the BCVA of the web-based and manifest assessment <sup>a</sup> |  |  | BCVA in logMAR of the web-based assessment |  |  | BCVA in logMAR of the manifest assessment |  |  |
|  | B | 95% confidence interval | P-value<sup>b</sup> | B | 95% confidence interval | P-value<sup>b</sup> | B | 95% confidence interval | P-value<sup>b</sup> |
| Sex<sup>c</sup> | 0.014 | -0.046 to 0.075 | 0.646 | -0.037 | -0.141 to 0.067 | 0.490 | -0.051 | -0.164 to 0.062 | 0.378 |
| Age | 0.000 | -0.002 to 0.002 | 0.870 | 0.003 | -0.001 to 0.007 | 0.132 | 0.003 | -0.001 to 0.007 | 0.160 |
| Use of mydriatics<sup>d</sup> | 0.032 | -0.044 to 0.107 | 0.409 | 0.145 | -0.201 to 0.491 | 0.412 | 0.113 | -0.174 to 0.401 | 0.441 |
| Visual acuity influencing comorbidities<sup>e</sup> | 0.012 | -0.047 to 0.071 | 0.695 | 0.056 | -0.072 to 0.184 | 0.393 | 0.044 | -0.081 to 0.169 | 0.492 |
| Symptoms<sup>f</sup> | 0.036 | -0.029 to 0.101 | 0.277 | 0.095 | -0.022 to 0.211 | 0.110 | 0.059 | -0.067 to 0.184 | 0.358 |
| Interval<sup>g</sup> | -0.004 | -0.013 to 0.005 | 0.367 | -0.002 | -0.019 to 0.015 | 0.811 | 0.002 | -0.014 to 0.018 | 0.815 |
| B: beta value<br>a: absolute difference between BCVA calculated in logMAR (web-based minus manifest)<br>b: analyzed using an Generalized Estimating Equations to correct for inclusion of 2 eyes of one patient<br>c: reference female<br>d: reference no use of mydriatics<br>e: visual acuity influencing comorbidities such as: (secondary) cataract, keratitis, scleritis, corneal lesion or history of pars plana vitrectomy<br>f: reference no symptoms associated with uveitis (pain, floaters, photophobia, visual loss) at time of the manifest assessment<br>g: interval in days between the web-based assessment and manifest assessment |  |  |  |  |  |  |  |  |  |

